# Interactions between hippocampal activity and striatal dopamine in people at clinical high risk for psychosis: relationship to clinical outcomes

**DOI:** 10.1101/2020.12.08.20245902

**Authors:** Gemma Modinos, Anja Richter, Alice Egerton, Ilaria Bonoldi, Matilda Azis, Mathilde Antoniades, Matthijs Bossong, Nicolas Crossley, Jesus Perez, James M. Stone, Mattia Veronese, Fernando Zelaya, Anthony A Grace, Oliver D Howes, Paul Allen, Philip McGuire

## Abstract

**Background:** Preclinical models propose that the onset of psychosis involves increased hippocampal activity which drives subcortical dopaminergic dysfunction. We used multi-modal neuroimaging to examine the relationship between hippocampal regional cerebral blood flow (rCBF) and striatal dopamine synthesis capacity in people at clinical high risk (CHR) for psychosis, and investigated its association with subsequent clinical outcomes.

**Methods:** Ninety-five participants (67 CHR and 28 healthy controls) underwent pseudo-continuous arterial spin labelling and ^18^F-DOPA PET imaging at baseline. Clinical outcomes in CHR participants were determined after a median of 15 months follow-up, using the Comprehensive Assessment of At Risk Mental States (CAARMS) and the Global Assessment of Function (GAF) scale.

**Results:** CHR participants with a poor functional outcome (follow-up GAF<65, n=25) showed higher rCBF in the right hippocampus compared to CHRs with a good functional outcome (GAF≥65, n=25) (familywise error [FWE] p=0·026). The relationship between right hippocampal rCBF and striatal dopamine synthesis capacity was also significantly different between groups (pFWE=0·035); the association was negative in CHR with poor outcomes (pFWE=0·012), but non-significant in CHR with good outcomes. The correlation between rCBF in this right hippocampal region and striatal dopamine function also predicted a longitudinal increase in the severity of positive psychotic symptoms (p=0·041). The relationship between hippocampal rCBF and striatal dopamine did not differ in the total CHR group relative to controls.

**Interpretation:** These findings indicate that altered interactions between the hippocampus and the subcortical dopamine system are implicated in the pathophysiology of psychosis-related outcomes.

## Introduction

Two robust neurobiological findings in patients with psychosis are alterations in the structure and function of the hippocampus,^1-4^ and increased striatal dopamine synthesis capacity^5^. Moreover, research in people at clinical high-risk (CHR) for psychosis suggests that these findings are evident before the first episode of the disorder (^2,6-9^, except^10^). These human findings are consistent with data from preclinical studies, which also suggest that increased hippocampal activity may drive dopamine dysfunction through projections to the striatum.^11-13^ Contemporary models propose that this interaction plays a critical role in the onset of psychosis.^14^

To date, relatively few studies have examined the relationship between hippocampal activity and striatal dopamine function in patients, partly because it entails the combination of MRI and PET techniques in the same patient. Two studies that used functional MRI to assess task-related hippocampal activation and ^18^F-DOPA PET to measure striatal dopamine synthesis capacity reported the relationship between these measures in people at CHR for psychosis was significantly different to that in healthy controls.^15,16^ However, the patient samples were small, and this precluded investigation of whether changes in the hippocampal-striatal relationship were associated with subsequent clinical outcomes. In this study, we sought to address these issues by studying hippocampal activity and dopamine function in a larger CHR sample, and clinically monitoring the participants after scanning to determine their clinical outcomes. Because preclinical data particularly implicate increases in resting hippocampal activity (as opposed to task-related activation), we assessed hippocampal activity at rest by using pseudo-continuous arterial spin labeling (pCASL), which indexes resting activity by measuring regional cerebral blood flow (rCBF)^17^.

Participants who had recently presented with a CHR state and were largely medication-naÏve were studied using pCASL and ^18^F-DOPA PET to measure hippocampal rCBF and striatal dopamine synthesis capacity, respectively. After the two baseline scans had been completed, participants were followed clinically to determine their clinical outcomes. We tested the hypothesis that within the CHR sample, clinical outcomes would be related to the nature of the association between hippocampal rCBF and striatal dopamine synthesis capacity at baseline.

## Methods

### Participants

A total of 95 individuals were examined, comprising participants from two independent studies conducted at King’s College London (appendix, pp 7-8 shows baseline sample characteristics by dataset). Both studies used the same clinical and neuroimaging methods. Ethical approval was obtained from the National Health Service UK Research Ethics Committee, and all participants provided written informed consent to participate.

CHR participants (n=67) were recruited from two early detection services: OASIS (Outreach and Support in South London^18^), part of the South London and Maudsley NHS Trust; and CAMEO (Cambridge Early Onset service), part of the Cambridge and Peterborough NHS Trust. Inclusion criteria were: (i) meeting operationalized criteria for CHR for psychosis, as determined with the Comprehensive Assessment of At-Risk Mental States (CAARMS^19^); (ii) no current/past diagnosis of psychotic/neurological disorder assessed with the structured clinical interview for diagnosis (SCID^20^); (iii) no substance abuse or dependence according to DSM-V criteria;^21^ and (iv) no contraindication to MRI or PET scanning.

Healthy controls (HC, n=28) were recruited from the same geographical area and met the following inclusion criteria: no personal/familial history of psychiatric/neurological disorder assessed using the SCID;^20^ no use of prescription medication as assessed via self-report; no substance abuse/dependence according to DSM-V criteria;^21^ and no contraindication to MRI or PET scanning.

### Clinical and Functional Measures

#### Baseline

All participants were assessed with the National Adult Reading Scale (NART^22^) to estimate premorbid IQ, and the Global Assessment of Functioning (GAF^23^) scale to measure overall social and occupational functioning. Participants provided information on cannabis use (yes/no). In all CHR participants, the severity of psychotic symptoms was evaluated using the CAARMS.^19^

#### Follow-up

Subsequent to baseline, CHR participants were clinically monitored in the community by an early detection team, which provided practical and psychological support. Clinical outcomes were assessed in face-to-face interviews after a median of 14·8 (interquartile range=11·1–22·1) months. The change in severity of positive psychotic symptoms between baseline and follow-up was calculated as follows:^10^

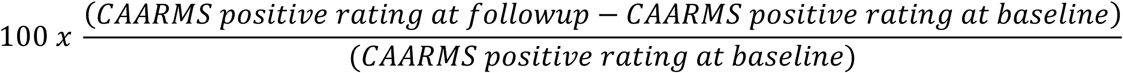

To examine overall functional outcome, following previous studies^24-26^ the CHR sample was dichotomized according to the GAF score at follow-up, with scores ≥65 defined as ‘good’, and scores <65 defined as ‘poor’. Transition to frank psychosis was defined using the criteria in the CAARMS,^19^ with the diagnosis confirmed using the SCID.^20^

### pCASL Acquisition and Preprocessing

The pCASL acquisition parameters were identical for both datasets, both acquired using the same General Electric Signa HDX 3.0T scanner at the Centre for Neuroimaging Sciences, King’s College London. The image acquisition parameters and preprocessing steps were identical for both datasets, are described in detail in our previous publications from these two ASL datasets,^6,7,27^ and summarized in the appendix (p 3). The resulting smoothed, normalized individual CBF maps were used for analysis.

### PET Acquisition and Preprocessing

PET imaging data were obtained on a Siemens Biograph 6 HiRez PET scanner (Erlangen, Germany) for one dataset and a GE Healthcare system (Chicago, Illinois) for the second dataset, in 3D mode. The PET data acquisition and preprocessing procedures are explained in detail in our previous publications from these two PET datasets,^10,28^ and summarized in the appendix (p 3). Our primary measure was the whole striatal influx constant (K_*i*_^cer^, min^-1^). Time-activity curves were visually inspected and K_*i*_^cer^ was calculated using the Patlak–Gjedde graphical approach adapted for a reference tissue input function.^29^ This approach has previously been shown to have good reliability, with intraclass correlation coefficients for the whole striatum of over 0·8.^30^

### PET Data Harmonization

Because the PET data were acquired with two different scanners, we used ComBat^31-33^ to harmonize the respective PET datasets. The ComBat algorithm successfully removes unwanted variation induced by scanner differences, while preserving biological variability between individuals by using an empirical Bayes framework.^31-33^ Two advantages of this approach over other methods are that it improves the removal of scanner effects in datasets with small sample sizes, and does not make any assumptions about the neuroimaging technique being used.^31,34^ The ComBat software was accessed from GitHub (https://github.com/Jfortin1/ComBatHarmonization) and the harmonization algorithm was performed in *R* version 3.6.0. In order to preserve between-subject variability, the analysis included age, sex, outcome group, CAARMS at baseline and at follow-up, GAF at baseline and at follow-up as covariates. Given preclinical and clinical evidence of a potential effect of cannabis use on dopamine synthesis and release,^35-37^ current cannabis use was also included as covariate in the ComBat analysis. By doing this, the algorithm minimized any differences between the two datasets that were not explained by any of these variables.

### Statistical Analysis

Clinico-demographic variables were compared using independent *t*-tests for continuous variables, and chi-square tests for categorical variables using the Statistical Package for Social Sciences (SPSS) version 26 (Chicago, IL). Significance was set at p<0·05 (two-tailed).

#### Global CBF

To exclude potential group differences in global CBF, global CBF values were extracted from each participant and subjected to independent t-tests in SPSS. The Automatic Software for ASL Processing (ASAP) 2.0 toolbox^38^ was used to extract average CBF values from the ICBM-152 mask as obtained from the DARTEL toolbox in SPM, and thresholded to contain voxels with a >0·20 probability of being grey matter.

#### Functional outcomes

The relationship between hippocampal rCBF and whole striatal dopamine synthesis capacity by functional outcome group was examined using SPM12, dividing the CHR sample into two groups at follow-up: a good functional outcome group (CHR-Good; GAF≥65) and a poor functional outcome group (CHR-Poor; GAF<65). Individual K_*i*_^cer^ values were entered as regressors in a voxel-wise ANCOVA to examine group differences in the relationship between whole striatal dopamine synthesis capacity and hippocampal rCBF in CHR-Poor compared to CHR-Good individuals, including age, sex and mean global CBF as covariates of no interest. Effects were considered significant at a voxel-wise height threshold of family-wise error (FWE) p<0·05 after small volume correction for region-of-interest analyses using a pre-specified anatomical mask of the bilateral hippocampus, derived from the WFU_Pickatlas toolbox (appendix, p 11). We also investigated whether the relationship between hippocampal rCBF and striatal dopamine synthesis capacity in the total CHR sample differed from HCs using an analogous approach in SPM12 as described above for the CHR subgroups.

#### Change in psychotic symptoms

The relationship between whole striatal dopamine synthesis capacity and hippocampal rCBF at baseline and subsequent changes in the severity of positive symptoms was investigated using linear regression in SPSS. Individual values from significant loci of interaction (cluster average) between striatal dopamine and hippocampal rCBF were extracted from the above ANCOVA using the MarsBar toolbox^39^ in SPM12 and then subjected to linear regression in SPSS, analyzing the relationship to percent change in CAARMS positive symptoms in the CHR sample (p<0·05).

#### Exploratory analyses

Additional exploratory analyses were conducted using measures of dopamine function in striatal subdivisions (limbic, associative and sensorimotor^40^). We assessed (i) group differences in dopamine function by striatal subdivision using a multivariate GLM in SPSS, and (ii) and group x dopamine synthesis capacity by striatal subdivision x hippocampal rCBF in SPM12, using the same procedures as in the main interaction analysis above. In view of evidence that perfusion abnormalities in CHR individuals may be particularly marked in the CA1 subregion of the hippocampus,^1,2,41^ we also examined the relationship between whole striatal dopamine synthesis capacity and rCBF using separate masks for the bilateral CA1, CA2, CA3, dentate gyrus, and subiculum in SPM12 (appendix, pp. 3, 12). As above, all SPM12 analysis used a significance threshold of pFWE<0·05.

## Results

All 95 participants received both PET and pCASL scans. There were no significant differences between the CHR and HC groups in age, sex, estimated premorbid IQ or cannabis use. As expected, CHRs had lower levels of overall functioning (GAF score) compared to HCs (Table). Most CHR participants were naÏve to antipsychotic medication (93%), and antidepressant-free (70%). Of the 67 CHR participants, 50 were followed-up clinically, while 17 were lost to follow-up. A comparison between these participants showed no significant differences in clinico-demographic variables (appendix, p 9).

**Table.** Baseline sample characteristics.

**Table. Baseline sample characteristics.**
|  | HC<br>(n=28) | CHR<br>(n=67) | HC vs CHR |  | CHR-Good<br>(n=25) | CHR-Poor<br>(n=25) | CHR-Good vs<br>CHR-Poor |  |
| --- | --- | --- | --- | --- | --- | --- | --- | --- |
| | | | *T/F or*<br>$\chi^2$ | *P* | | | *T/F or*<br>$\chi^2$ | *P* |
| Age in years, mean (SD) | 24.43<br>(4.43) | 22.80<br>(4.23) | 1.70 | 0.09 | 22.20<br>(4.53) | 23.74<br>(3.96) | -1.28 | 0.21 |
| Sex (male), N | 16<br>(57.1%) | 40<br>(59.7%) | 0.05 | 0.82 | 15 (60%) | 15 (60%) | 0.000 | 1.00 |
| Years of education, mean (SD) | 15.11<br>(3.07) | 13.98<br>(2.43) | 1.91 | 0.06 | 13.96<br>(2.35) | 13.68<br>(2.64) | 0.40 | 0.69 |
| Premorbid IQ estimate | 103.12<br>(11.70) | 106.00<br>(10.75) | -1.13 | 0.26 | 105.89<br>(10.10) | 107.93<br>(11.01) | -0.67 | 0.51 |
| GAF, mean (SD) | 88.13<br>(8.64) | 58.31<br>(9.27) | 14.55 | <0.001 | 58.36<br>(10.37) | 56.92<br>(9.07) | 0.52 | 0.60 |
| CAARMS Total, mean (SD) | NA | 41.53<br>(18.32) | NA | NA | 38.71<br>(16.45) | 43.29<br>(19.55) | -.088 | 0.39 |
| CAARMS Positive, mean (SD) | NA | 8.00<br>(3.58) | NA | NA | 7.44<br>(3.86) | 8.64 (3.66) | -1.13 | 0.27 |
| CAARMS Negative, mean (SD) | NA | 5.89<br>(3.65) | NA | NA | 6.04<br>(3.91) | 5.75 (3.51) | 0.27 | 0.79 |
| Antipsychotics, N | NA | 5 | NA | NA | 3 | 0 | 3.19 | 0.07 |
| Antidepressants, N | NA | 20 | NA | NA | 8 | 8 | 0.000 | 1.00 |
| Cannabis use, N | 15<br>(53.6%) | 34<br>(50.7%) | 0.18 | 0.67 | 14 (56.0) | 10 (40%) | 1.28 | 0.26 |
| Global rCBF, mean (SD) | 277.86<br>(45.68) | 268.57<br>(54.39) | 0.63 | 0.43 | 271.64<br>(52.15) | 261.50<br>(61.46) | 0.40 | 0.53 |
| Striatal $K_i^{cer}$, mean (SD) | 0.0126<br>(0.001) | 0.0128<br>(0.001) | 0.82 | 0.37 | 0.0131<br>(0.001) | 0.0127<br>(0.001) | 1.93 | 0.17 |

**Abbreviations:** CAARMS, Comprehensive Assessment of At Risk Mental State; CHR, clinical high risk; GAF, Global Assessment of Function; SD, standard deviation.

Of the 50 CHR individuals with available clinical follow-up, 25 had a good functional outcome (CHR-Good), and 25 had a poor functional outcome (CHR-Poor) (appendix, p 13). No significant differences were found in clinico-demographic variables (including medication or cannabis use) at baseline between these groups (Table). At follow-up, the majority of these 50 CHR participants remained antipsychotic-(90%) and antidepressant-free (78%). Six individuals of the total CHR sample developed a psychotic disorder during the follow-up period. There was no significant difference between the functional outcome subgroups in the proportion of participants who had transitioned to psychosis in each (2/25 in CHR-Good, 4/25 in CHR-Poor; χ^2^=0·758, p=0·384).

### Functional outcomes

We observed a significant interaction between rCBF, whole striatal dopamine synthesis capacity and group (CHR-Good vs CHR-Poor) in the right hippocampus (xyz: 40, −12, −24; k=14; t=3·56, z=3·32, pFWE=0·035, Figure 1A). This effect was driven by a significant negative association in the CHR-Poor subgroup (xyz: 38, −8, −24; k=20; t=3·99, z=3·66, pFWE=0·012), which was absent in the CHR-Good subgroup (pFWE>0·05) (Figure 1B). Excluding the three CHR participants who had been treated with antipsychotics from the analysis did not change the results (xyz: 38, −8, −24; k=17; t=3·93, z=3·60, pFWE=0·015).

**Figure 1.**
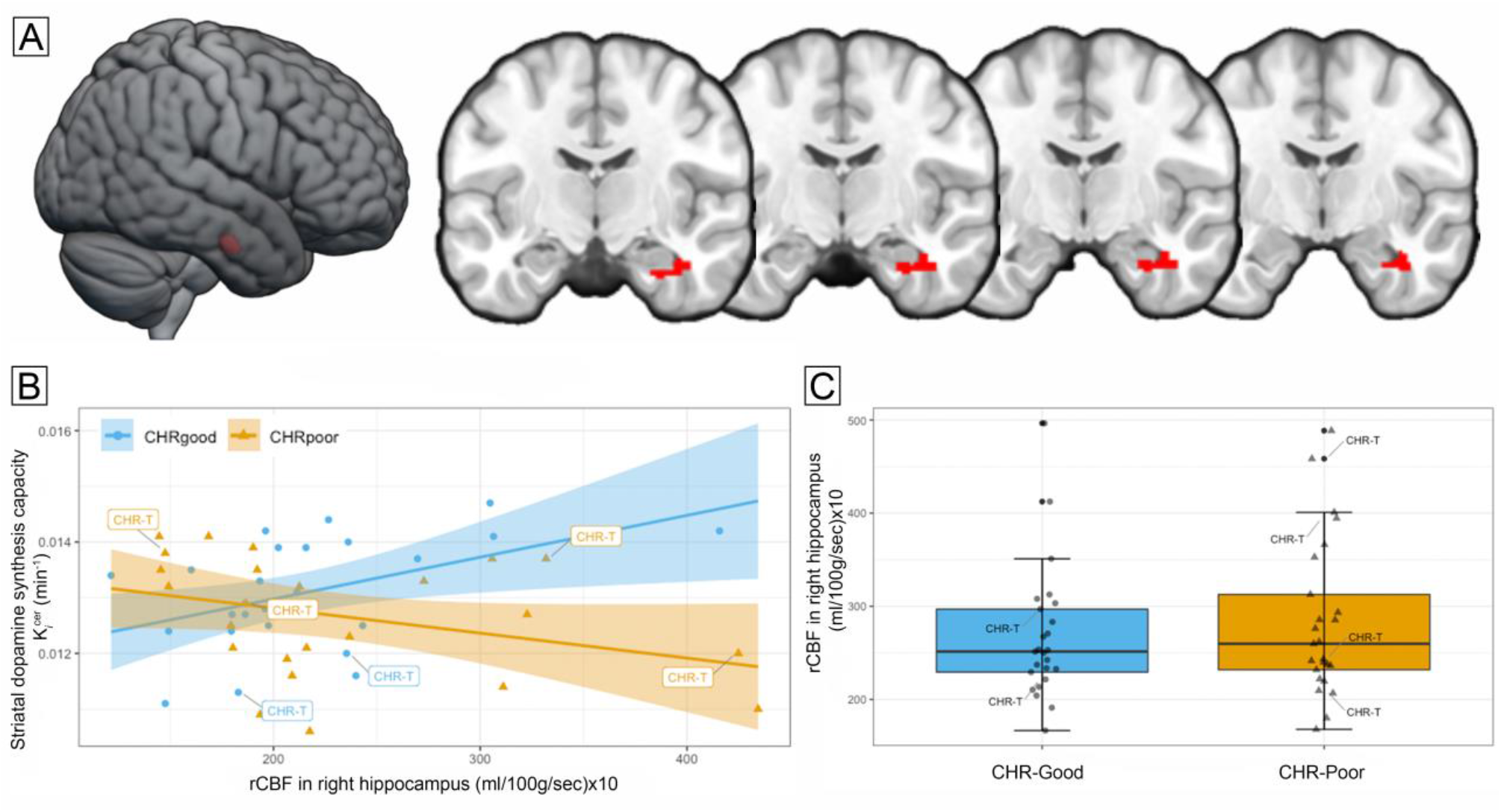
Relationship between hippocampal activity, striatal dopamine synthesis capacity and functional outcomes. (A) Significant group (CHR-Poor vs CHR-Good) x rCBF x striatal dopamine interaction in the right hippocampus (pFWE=0·035), overlaid on a standard brain template. (B) Scatterplot depicts within-group associations between hippocampal rCBF and striatal dopamine (CHR-Good, blue circles, pFWE>0·05; CHR-Poor, orange triangles, pFWE=0·015), with regression lines and 95% CIs. (C) Boxplots show increased rCBF in this right hippocampal cluster in CHR-Poor individuals compared to CHR-Good (pFWE=0·026). CHR-T, clinical high-risk individuals who subsequently transitioned to psychosis.

Analyzing the data separately by imaging modality revealed that individuals in the CHR-Poor group had significantly higher rCBF in this right hippocampal region compared to the CHR-Good group (xyz: 40, −12, −24; k=17; t=3·68, z=3·42, pFWE=0·026, Figure 1C). No differences were found in either global rCBF (F[48]=0·395, p=0·532), dopamine synthesis capacity in the whole striatum (F[48]=1·933; p=0·171), or by subdivisions (appendix, pp 10, 14).

The total CHR and HC groups did not differ in terms of global rCBF (F[93]=0·824, p=0·366), total hippocampal rCBF or by subfield (pFWE>0·05), dopamine synthesis capacity in the whole striatum (F[94]=0·824; p=0·366) or by subdivisions (appendix, pp. 10, 15). No significant interactions were found between hippocampal rCBF, striatal dopamine and baseline group status (total CHR vs HC; pFWE>0·05).

### Change in psychotic symptoms

We next examined the relationship between the hippocampal-striatal interaction and the subsequent worsening of positive psychotic symptoms. The model showed a direct relationship between the individual values extracted from the significant cluster of hippocampal rCBF x whole striatal dopamine interaction and percent change in positive psychotic symptoms (β=0·296, R^2^=0·087, df=47, p=0·041) (Figure 2). This indicated that the stronger the association between striatal dopamine and hippocampal rCBF at baseline, the greater the worsening of symptoms over the subsequent follow-up period. This effect remained evident as a strong trend after CHR participants who had been treated with antipsychotics (n=3) were excluded from the analysis (β=0·288, R^2^=0·083, df=44, p=0·055).

**Figure 2.**
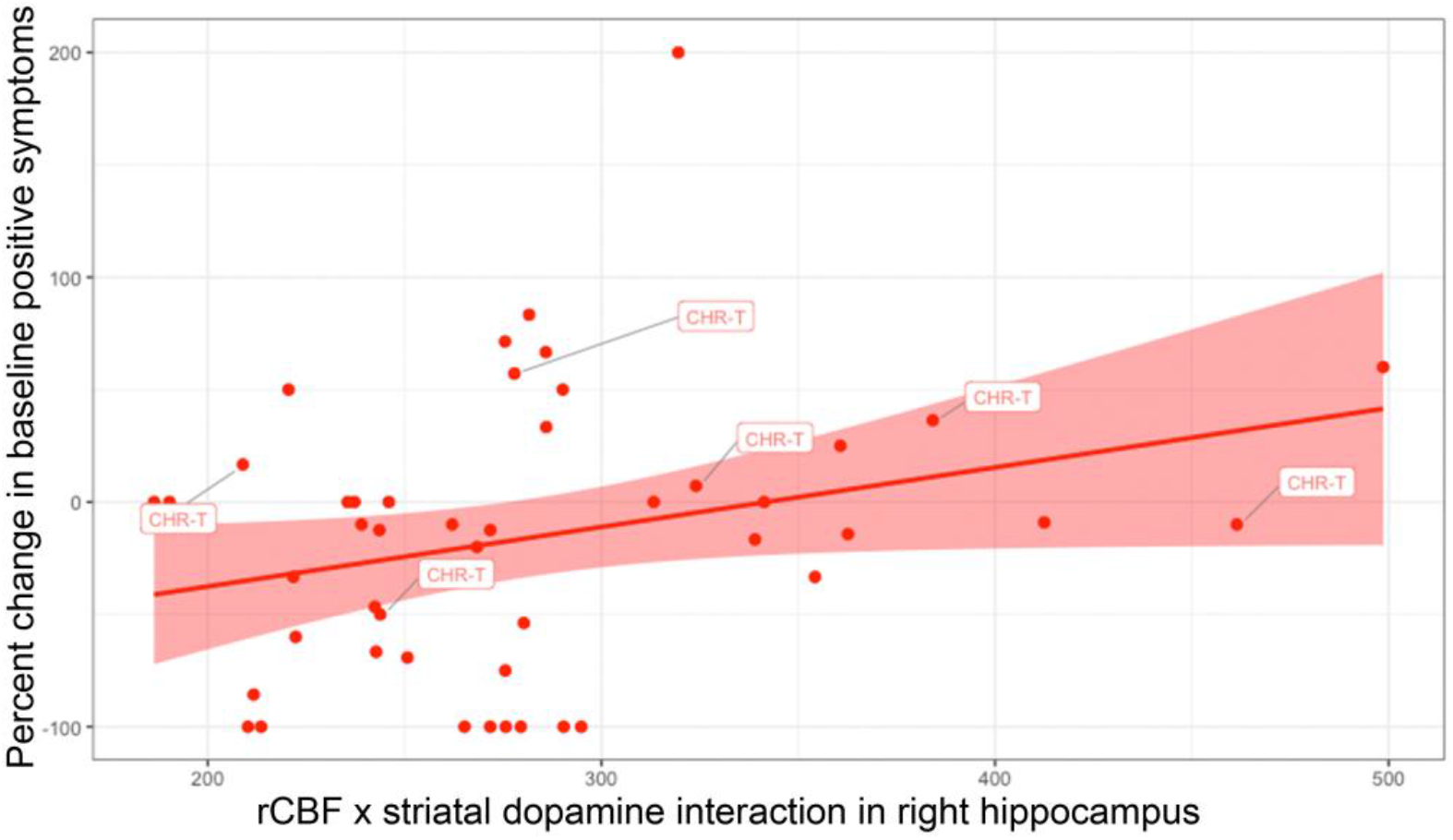
The relationship between hippocampal activity and striatal dopamine function predicted the subsequent worsening of positive psychotic symptoms. The scatterplot depicts a linear regression in which percent change in baseline positive symptoms is the dependent variable and the cluster-averaged values of the rCBF x K_*i*_ interaction is the independent variable, with regression lines and 95% CIs (p=0·041). CHR-T, clinical high-risk individuals who subsequently transitioned to psychosis.

### Exploratory analyses

Exploratory analyses using measures of dopamine synthesis capacity from striatal subdivisions also identified significant group interactions for the associative and the sensorimotor subdivisions. In both cases, these were driven by negative associations with total hippocampal rCBF in the CHR-Poor subgroup (appendix, pp. 5, 16). Repeating the main analyses using rCBF measures from hippocampal subfields revealed a significant group interaction with whole striatal dopamine for the right CA1. Similar to the finding for total hippocampal rCBF, this reflected a negative association in the CHR-Poor subgroup that was not evident in the CHR-Good subgroup (appendix, p 5). Furthermore, separate rCBF-only group comparisons by hippocampal subfields showed greater rCBF in the right CA1 in CHR-Poor compared to CHR-Good (appendix, pp. 3, 5). Finally, supplemental analysis with other outcome definitions (psychosis transition/non-transition; CHR remission/non-remission^42^) are reported for illustration purposes in the appendix (appendix, pp. 4, 5).

## Discussion

Our main finding was that functional outcome in CHR individuals was related to the nature of the association between increased rCBF in the right hippocampus and striatal dopamine synthesis capacity at baseline. Specifically, a poor functional outcome was linked to elevated rCBF in the right hippocampus, and to a negative association between rCBF in this region and striatal dopamine synthesis capacity, compared to CHRs with a good outcome. Furthermore, the relationship between rCBF in this right hippocampal region and striatal dopamine function was also linked to a worsening of psychotic symptoms subsequent to scanning.

Preclinical studies demonstrated that hippocampal hyperactivity leads to striatal hyperdopaminergia and psychosis-like behaviors, such as increased amphetamine-induced locomotion.^11-13^ Moreover, pharmacological^43,44^ and chemogenetic^45^ manipulations that normalize hippocampal hyperactivity can correct aberrant dopamine neuron population activity in the striatum.^44^ Neuroimaging studies in people at CHR for psychosis reported both increased hippocampal rCBF and hippocampal blood volume compared to healthy controls,^2,6,7^ while functional MRI studies found attenuated hippocampal responses in CHR during memory^15^ and salience processing^9^ tasks. Furthermore, increased hippocampal blood volume at baseline was linked to transition to psychosis,^2^ and the longitudinal normalization of hippocampal rCBF was associated with remission from the CHR state.^6^ Parallel work using PET reported elevated striatal dopamine synthesis capacity in CHR individuals compared to healthy controls (^28,46,47^, except^10^), linked to the subsequent onset of psychosis^48,49^, and to a worsening of psychotic symptoms.^10^ While there is thus independent evidence for both altered hippocampal function and striatal hyperdopaminergia in the CHR state, the relationship between them is less clear. Two studies reported that the correlation between task-related hippocampal activation and striatal dopamine in CHR individuals was significantly different to that in healthy controls, being negative as opposed to positive.^15,16^ These were cross-sectional studies in small CHR samples, precluding investigation of the extent to which changes in the hippocampal-striatal relationship are linked to subsequent clinical outcomes. Our study addressed this issue by studying a larger sample of CHR individuals with both pCASL and ^18^F-DOPA PET, and following them up after scanning to determine their clinical outcomes. Our main hypothesis, that an altered relationship between hippocampal rCBF and striatal dopamine function would be associated with adverse clinical outcomes, was confirmed: a negative association between heightened hippocampal rCBF and striatal dopamine was linked to a poor functional outcome and to an increase in the severity of psychotic symptoms. Moreover, exploratory analyses of hippocampal subfields indicated that these findings were particularly evident in the CA1 subregion, and in the associative and sensorimotor subdivisions of the striatum, supporting evidence implicating the CA1 and the associative striatum as key foci of dysfunction in psychosis.^2,5,41,50,51^

The direction of the altered relationship between hippocampal rCBF and striatal dopamine function in CHR-Poor individuals was negative. Given the evidence that both hippocampal activity and dopamine function are elevated in CHR individuals, and the notion that one drives the other, one might have expected adverse outcomes to be linked to a *positive* correlation between these measures. Our findings indicate that, in CHR individuals with a poor functional outcome, those with greater hippocampal rCBF showed lower striatal dopamine synthesis capacity. One possibility is that these findings may challenge the main assumption of a causal relationship between elevated hippocampal activity and striatal dopamine in psychosis. Rodent studies have shown a clear positive relationship between hippocampal activity and dopaminergic neuron function in a system that operates at equilibrium.^52,53^ Hence, it is also possible that the observed negative correlation reflects a break-down of this equilibrium in CHR individuals with adverse clinical outcomes, as this in turn was found to significantly predict the worsening of positive symptoms over time. The relationship between hippocampal rCBF and striatal dopamine function may be non-linear or quadratic rather than linear, and it may also be indirect, involving additional subcortical regions such as the nucleus accumbens, ventral pallidum and the ventral tegmental area,^52,54^ as well as other neurotransmitter systems such as glutamate or GABA.^13,55^ Longitudinal studies with serial multimodal neuroimaging assays are warranted to establish the direction of causality.

In contrast with previous findings, no significant differences in striatal dopamine synthesis capacity between the total CHR group and HCs were found.^28,46,47^ This sample partially overlaps with a previous study from our group which reported a similar negative ^18^F-DOPA PET finding.^10^ We also did not find a significant difference in hippocampal rCBF between the CHR and HC groups, in contrast to our previous findings in larger studies that included the current cohorts.^6,7^ These differences may reflect the larger numbers of individuals who subsequently transitioned to psychosis during the follow-up period in those previous studies. The sample size imbalance (67 CHR vs 28 HC) may also weaken the statistical inferences about group differences for these analyses. However, this would not affect the results of our primary analyses based on functional outcomes involving two CHR subgroups of equal size. Overall, our findings extend prior single-modality CHR studies indicating that the altered relationship between hippocampal rCBF and striatal dopamine synthesis capacity may be specific to CHR individuals with poor functional outcomes, and could relate to the development of psychopathology other than psychosis later in life (e.g., affective disorders) in these individuals.^56,57^ Further studies with longer longitudinal follow-up and a wider assessment of psychopathology outcomes are required to expand on the mechanisms underlying poor functional outcomes in the CHR state.

Our findings should be considered in light of some limitations. In order to form as large a sample of CHR individuals with pCASL, PET and complete follow-up data as possible, we combined two independent datasets. Participants in these two studies were recruited from the same centers, and the clinical and pCASL measures were identical. Although the PET data in the two studies were acquired at the same center using the same methods, the scanner models were different. We controlled for effects of the latter using ComBAT, a robust method to remove unwanted technical variability induced by scanner differences.^33^ Because previous clinical follow-up of the two CHR samples comprising the total cohort indicated that only a small number of participants had made a transition to psychosis (n=6) or remitted from the CHR state (n=16), there would not have been sufficient power to detect interactions between the baseline neuroimaging measures (appendix, pp. 4-5). We therefore used level of functioning as our primary outcome, comparing subgroups with ‘good’ and ‘poor’ GAF follow-up scores. This approach permits inclusion of the entire sample, and yields subgroups of approximately equal size.^24-26^ There was no significant difference between the functional outcome subgroups with regard to the number of individuals transitioning to psychosis in each, but such difference may be hard to find given that only six CHR individuals transitioned. Future studies may explore associations with transition to psychosis by conducting multi-modal neuroimaging and follow-up in a larger CHR sample, although this would be logistically demanding, and would entail a multi-center design. A strength of the study was that almost all of the CHR individuals were naÏve to antipsychotic medication, minimizing the likelihood of confounding effects on the findings. Moreover, exclusion of the small number of participants who had been treated with antipsychotics did not alter the main findings.

In conclusion, our findings suggest that adverse clinical outcomes in the CHR state are linked to interactions between heightened resting hippocampal activity and striatal dopamine function, and support future research to examine the effect of stabilizing hippocampal hyperactivity to prevent the development of psychosis-related outcomes.^14^

## Data Availability

We are unable to share data because participants did not provide consent for data sharing.

## Contributors

GM, AE, IB, MA, JP, MA and MB contributed to data acquisition. GM, AR, AE, IB, NC, FZ and MV contributed to data analysis. GM and AR wrote the initial draft of the manuscript. ODH, JS, PA, AAG, and PM obtained funding, and were involved in the initial design of the research. All authors contributed to data interpretation and submission of the final manuscript.

## Acknowledgments

This work was supported by a Wellcome Trust Programme Grant to PM, PA, ODH, JS & AAG (grant number 091667, 2011), and an MRC Research Grant to PM (grant number G0700995). GM is supported by a Sir Henry Dale Fellowship jointly funded by the Wellcome Trust and the Royal Society (grant number 202397/Z/16/Z). We thank the MRI radiographers for their expert assistance, the study volunteers for their participation, and we gratefully thank members of the OASIS and CAMEO clinical teams who were involved in the recruitment and management of the CHR participants in this study.

