## appendix for "Interactions between hippocampal activity and striatal dopamine in people at clinical high risk for psychosis: relationship to clinical outcomes"

### Supplemental Methods

#### pCASL acquisition and preprocessing

Parameters for data acquisition were identical for both datasets. Subjects were scanned at a General Electric Signa HDX 3·0T scanner with their eyes open, using an 8-channel phased array head coil at the Department of Neuroimaging, Institute of Psychiatry, Psychology and Neuroscience. A high-resolution T2-weighted Fast Spin Echo (FSE) image (TE=54·58ms, TR=4380ms, flip angle 90°, field of view=240) was acquired for image registration.

Pseudo-Continuous Arterial Spin Labelling (pCASL) scans were acquired with a 3D FSE spiral multi-shot readout, after a post-labelling delay of 1·5 s. Parameters of the image readout were as follows: TE=32 ms, TR=5500 ms, spatial resolution of 1x1 mm in plane, 60 slices of 3 mm thickness and three pairs of tagged-untagged images were collected. To maximize the sensitivity to blood perfusion, suppression of the background static tissue signal was achieved by selective saturation of the image slab at 4·3s before acquisition, selective inversion 3 s before acquisition and non-selective inversions at 1·5 s, 764 ms, 334 ms and 84 ms before imaging. Calibration images were collected with the same imaging sequence and were used to quantify blood flow in physiological units (ml blood/100 gm tissue/min).

The sensitivity of the image to water was calibrated at each voxel.[1-3] A low-resolution sensitivity map was created by using a neighbourhood maximum algorithm to avoid regions with partial volume of suppressed fluid. With this co-registered sensitivity map C, cerebral blood flow (CBF) was calculated following the guidelines reported by Alsop et al.[2] The whole ASL pulse sequence, including the acquisition of calibration images, was performed in 6:08 minutes.

FSL (FMRIB Software Library, http://www.fmrib.ox.a.c.uk/fsl) and Statistical Parametric Mapping (SPM12; http://www.fil.ion.ucl.ac.uk/spm/) were used to preprocess CBF maps. The following steps were applied: (1) extra-cerebral signal was removed from the T2 scan using the "Brain Extraction Tool" (BET) of FSL28 and the resulting skull-stripped T2 volume and its corresponding T2 binary mask were then co-registered to the ASL scan; (2) extra-cerebral signal was then eliminated from the CBF map by multiplying it with the co-registered binary brain mask; (3) the skull-stripped CBF map was co-registered back to the space of the original T2 scan to return to their original frame of reference; (4) the T2 and the multiplied ASL scans (step 2) were normalized to SPM's T2 template; (5) the normalized individual CBF map was spatially smoothed using a 6mm Gaussian smoothing kernel.

For more information, see our recently published papers with this methodology.[4-6]

#### PET acquisition and preprocessing

Participants were asked to refrain from eating, drinking (except water), and alcohol consumption for 12 h prior to the scan, and from smoking for 2 h prior to the scan.[7] One hour before the scan, all participants received 400 mg of entacapone, a peripheral catechol-*o*-methyl-transferase inhibitor, and 150 mg of carbidopa, a peripheral aromatic acid decarboxylase inhibitor. Participants were positioned in the scanner with the orbitomeatal line parallel to the transaxial plane of the tomograph. To correct for attenuation, the head position was marked and monitored, and a CT scan was conducted. Thirty seconds after the start of the dynamic PET scan, approximately 150 MBq of $^{18}$F-DOPA was administered by bolus intravenous injection. PET data were acquired in 32 frames of increasing duration and the whole scan was performed in 95 min (frame intervals: 8x15, 3x60, 5x120, 16x300s).

In terms of image preprocessing, a mutual information algorithm was used to correct for head movement.[8] SPM8 was used to automatically normalise a tracer-specific $^{18}$F-DOPA template.[9] A striatal brain atlas using the definition described by Martinez et al. was applied, which includes dividing the striatum into a limbic, associative and sensorimotor subdivision, respectively, based on the predominant origin of projections to the striatum.[10]

#### Exploratory analysis of rCBF x whole striatal dopamine synthesis capacity by hippocampal subfields

Previous studies suggest that perfusion abnormalities in CHR individuals may be particularly evident in the CA1 subregion of the hippocampus.[11-13] For completeness, we examined the relationship between hippocampal rCBF and striatal dopamine synthesis capacity in the CHR-Good compared to CHR-Poor subgroup in SPM12, using analogous procedures to those described for our main analysis. Separate masks for bilateral CA1, CA2, CA3, dentate gyrus, and subiculum were anatomically defined using the cytoarchitectonic probabilistic atlas[14] in the SPM Anatomy toolbox[15] (Figure S2) and used for small volume correction (SVC) in an ANCOVA in SPM12 with the rCBF maps by functional outcome subgroup, individual dopamine ($K_i^{cer}$) values as regressors, and including age, sex and mean global CBF as covariates of no interest. For completeness, in this same design we also examined whether rCBF between the CHR-Good and CHR-Poor groups differed by hippocampal subfield. As in our main analyses, we used a voxel-wise height threshold of family-wise error (FWE) correction of $p<0·05$ after small volume correction (SVC) with the pre-defined hippocampal ROI mask.

**Supplementary analysis of other CHR clinical outcomes**

*Transition / non-transition to psychosis*

The relationship between hippocampal rCBF and whole striatal dopamine synthesis capacity ($K_i^{cer}$, $min^{-1}$) by psychosis transition/non-transition outcome groups was examined using SPM12, dividing the CHR sample into two groups: CHR individuals who developed a psychotic disorder during follow-up period (CHR-T), and those who did not (non-transitioned, CHR-NT). Individual $K_i^{cer}$ values were entered as regressors in a voxel-wise ANCOVA to examine between- and within-group associations of whole striatal dopamine synthesis capacity with hippocampal rCBF in CHR-T compared to CHR-NT individuals. Age, sex and mean global CBF were included as covariates of no interest in the ANCOVA. In addition, exploratory analyses were conducted using measures of dopamine function in striatal subdivisions (limbic, associative and sensorimotor). First, we assessed group differences in dopamine function by striatal subdivision using a multivariate GLM in SPSS. Second, we assessed group interactions on the relationship between dopamine synthesis capacity by striatal subdivision and hippocampal rCBF in SPM12, using the same procedures as in the main interaction analysis. Third, we tested the relationship between hippocampal rCBF and dopamine synthesis capacity in SPM12 using separate masks for the bilateral CA1, CA2, CA3, dentate gyrus, and subiculum. A significance threshold of $pFWE<0·05$ was used for all SPM12 analyses.

*CHR remission / non-remission*

Based on previous research, we also analyzed for completeness the relationship between hippocampal rCBF and whole striatal dopamine synthesis capacity ($K_i^{cer}$, $min^{-1}$) by remission/non-remission outcome groups. This was examined in SPM12, by allocating CHR subjects to either remission (CHR-R, no longer meeting CHR criteria with the Comprehensive Assessment of At-Risk Mental States (CAARMS)[16]) or non-remission (CHR-NR, meeting CHR or psychosis criteria) groups on follow-up assessment. We used the same procedures as in the main hippocampal rCBF x striatal dopamine synthesis capacity ($K_i^{cer}$, $min^{-1}$) interaction analysis by functional outcome subgroups and above for CHR-T vs CHR-NT: individual $K_i^{cer}$ values were entered as regressors in a voxel-wise ANCOVA to examine between- and within-group associations of whole striatal dopamine synthesis capacity with hippocampal rCBF in CHR-R compared to CHR-NR individuals, including age, sex and mean global CBF as covariates of no interest. In additional exploratory analyses, group differences in (1) dopamine function by striatal subdivision were investigated using a multivariate GLM in SPSS, (2) group interactions on the relationship between dopamine synthesis capacity by striatal subdivision and hippocampal rCBF were examined in SPM12, (3) group interactions on the relationship between whole striatal dopamine synthesis capacity rCBF by hippocampal subfields (bilateral CA1, CA2, CA3, dentate gyrus, and subiculum). A significance threshold of $pFWE<0·05$ was used for all SPM12 analyses.

### Supplemental Results

#### Relationship between hippocampal rCBF and dopamine synthesis capacity by striatal subdivision

Within the CHR-Poor group, hippocampal rCBF was negatively associated with dopamine synthesis capacity in the associative (xyz: 38, -8, -24; k=27; t=4·32, z=3·92, pFWE=0·005) and limbic striatum (xyz: 28, -6, -28; k=59; t=3·67, z=3·40, pFWE=0·027), but only at trend level in the sensorimotor striatum (xyz: 38, -8, -24; k=2; t=3·11, z=2·93, pFWE=0·099). These associations were absent in the CHR-Good group. Group interaction analyses showed that the relationship between hippocampal rCBF and dopamine synthesis capacity in the associative striatum was significantly different between CHR groups (xyz: 40, -12, -24; k=8; t=3·45, z=3·22, pFWE=0·046), as well as in the sensorimotor striatum (xyz: 40, -12, -24; k=9; t=3·55, z=3·31, pFWE=0·036), while the group interaction was evident as a strong trend in the limbic striatum (xyz: 34, -10, -26; k=53; t=3·38, z=3·16, pFWE=0·054) (Figure S5).

#### Exploratory analysis of rCBF x whole striatal dopamine synthesis capacity by hippocampal subfields

Analysis by hippocampal subfields revealed that the interaction between whole striatal dopamine synthesis capacity, group (CHR-Good vs CHR-Poor) and hippocampal rCBF was significant in the right CA1 subregion (xyz 38, -12, -24, k=6, t=3·24, z=3·05, pFWE=0·042). This was driven by a significant negative association in CHR individuals with poor functional outcomes (xyz: 38, -12, -24; k=21; t=3·42, z=3·20, pFWE=0·027), which was absent in CHR individuals with good outcomes. No other significant effects were found in other subfields (CA2, CA3, dentate gyrus and subiculum).

Separate analyses of between-group differences in only rCBF, by hippocampal subfield showed elevated rCBF in the right CA1 subregion in CHR-Poor individuals compared to CHR-Good (xyz: 38, -12, -24; k=15; t=3·37, z=3·16, pFWE=0·031). No other significant effects were found in other subfields (CA2, CA3, dentate gyrus and subiculum).

#### Supplementary analyses of other CHR outcomes

<u>Transition / non-transition to psychosis</u>
Of the 50 CHR individuals with available clinical follow-up, six developed a psychotic disorder during follow-up period (CHR-T) and 44 did not (CHR-NT). No significant differences were found in demographic or clinical variables (including medication or cannabis use) at baseline between these groups. Of the 6 CHR-T individuals, five were naïve to antipsychotic or antidepressant medication (1 was taking antidepressants). Of the 44 CHR-NT individuals, 26 were naïve to antipsychotic or antidepressant medication (3 were taking antipsychotics, 15 antidepressants).

*Relationship between hippocampal rCBF and striatal dopamine*
There was no significant group interaction between rCBF, whole striatal dopamine synthesis capacity and group in the hippocampus (pFWE>0·05).

While there were no significant between-group differences in total hippocampal rCBF, there was a significant increase in rCBF in the left subiculum in CHR-T compared to CHR-NT (xyz: -24, -22, -26; k=17; t=3·21, z=3·03, pFWE=0·039). No other significant interaction by hippocampal subfield was found (pFWE>0·05). There were no significant differences between the two groups in striatal dopamine synthesis capacity in the whole striatum (F([48]=0·470; p=0·496; adjusted for age and sex F[46]=0·854, p=0·360) or by striatal subdivisions (Table S4).

*Exploratory analysis of hippocampal rCBF x dopamine synthesis capacity by striatal subdivision*
There was a significant group interaction between rCBF in the hippocampus and dopamine synthesis capacity in the associative striatum (xyz: -20, -22, -18; k=112; t=3·76, z=3·48, pFWE=0·021), which was driven by a positive association in the CHR-T subgroup (xyz: -20, -22, -18; k=128; t=3·89, z=3·58, pFWE=0·015). There were no significant interactions for any of the other subdivisions (limbic or sensorimotor, pFWE>0·05).

*Exploratory analysis of rCBF x whole striatal dopamine synthesis capacity by hippocampal subfields*
Analysis by hippocampal subfields showed a significant interaction between rCBF, whole striatal dopamine synthesis capacity and group (CHR-T vs CHR-NT) in the left subiculum (xyz: -22, -28, -16; k=87; t=3·28, z=3·08, pFWE=0·034), which was driven by a significant positive association in the CHR-T subgroup (xyz: -20, -24, -18; k=76; t=3·35, z=3·14, pFWE=0·029). There were no significant interactions with other hippocampal subfields (CA1, CA2, CA3, dentate gyrus, pFWE>0·05).

CHR remission / non-remission

Sixteen CHR individuals were in remission (CHR-R), and 33 individuals still met CHR criteria (non-remission, CHR-NR) at the follow-up assessment. There were no significant differences between CHR-R and CHR-NR subgroups in terms of age, sex, estimated premorbid IQ, cannabis use, and level of overall functioning. CHR individuals with persistent CHR status (CHR-NR) had significantly higher positive symptoms at baseline (mean: 9·21, SD=3·35) compared to CHR-R (mean: 5·94, SD: 3·62; df=47, p=0·003). Of the 16 CHR-R individuals, nine were naïve to antipsychotic or antidepressant medication (2 were taking antipsychotic medication, 5 antidepressant medication). Of the 33 CHR-NR individuals, 21 were naïve to antipsychotic or antidepressant medication (1 was taking antipsychotic medication, 11 antidepressant medication).

*Relationship between hippocampal rCBF and striatal dopamine*

There was no significant group interaction between rCBF, whole striatal dopamine synthesis capacity and group in the hippocampus (pFWE>0·05).

When analyzed separately, there were no significant differences between the CHR-R and CHR-NR subgroups in either total hippocampal rCBF or by hippocampal subfield (pFWE>0·05), striatal dopamine synthesis capacity in the whole striatum (F([47]=0·115; p=0·736; adjusted for age and sex F[45]=0·493, p=0·486) or by striatal subdivisions (Table S4).

*Exploratory analysis of hippocampal rCBF x dopamine synthesis capacity by striatal subdivision*

There were no significant group interactions between rCBF in the hippocampus and striatal dopamine synthesis capacity in any subdivisions (associative, limbic or sensorimotor, pFWE>0·05).

*Exploratory analysis of rCBF x whole striatal dopamine synthesis capacity by hippocampal subfields*

There were no significant group interactions between whole striatal dopamine synthesis capacity and rCBF in any hippocampal subfields (CA1, CA2, CA3, dentate gyrus, subiculum, pFWE>0·05).

### Supplemental Tables

**Table S1. Baseline characteristics of the first sample (NEUTOP).**

| | **HC (n=16)** | **CHR (n=45)** | **HC vs CHR** | | **CHR-Good (n=15)** | **CHR-Poor (n=19)** | **CHR-Good vs CHR-Poor** | |
|---|---|---|---|---|---|---|---|---|
| | | | **T or $\chi^2$** | **p** | | | **T or $\chi^2$** | **p** |
| Age in years, mean (SD) | 25·32 (4·46) | 22·92 (3·90) | 2·04 | 0·05 | 21·46 (2·93) | 23·92 (4·02) | -1·99 | 0·06 |
| Sex (male), N | 7 (43·8%) | 27 (60%) | 1·26 | 0·26 | 8 | 13 | 0·81 | 0·37 |
| Years of education, mean (SD) | 15·38 (3·96) | 14·39 (2·36) | 1·19 | 0·24 | 14·27 (2·38) | 14·11 (2·49) | 0·19 | 0·85 |
| Premorbid IQ estimate | 99·93 (12·35) | 105·84 (11·50) | -1·58 | 0·13 | 104·71 (9·82) | 107·61 (12·28) | -0·72 | 0·48 |
| GAF, mean (SD) | 91·84 (5·06) | 58·46 (9·10) | 17·99 | <0·001 | 56·93 (9·17) | 57·00 (9·64) | -0·02 | 0·98 |
| CAARMS Total, mean (SD) | n/a | 42·41 (20·68) | - | - | 42·73 (20·17) | 43·83 (21·92) | 0·63 | 0·88 |
| CAARMS Positive, mean (SD) | n/a | 8·66 (3·64) | - | - | 8·80 (3·93) | 8·79 (3·72) | 0·01 | 0·99 |
| CAARMS Negative, mean (SD) | n/a | 5·83 (3·91) | - | - | 6·53 (4·22) | 5·50 (3·67) | 0·75 | 0·46 |
| Antipsychotics, N | - | 4 (8·9%) | - | - | 2 (13·3%) | 0 | 2·69 | 0·10 |
| Antidepressants, N | - | 15 (33%) | - | - | 6 | 6 | 0·26 | 0·61 |
| Cannabis use, N | 7 | 19 | 0·01 | 0·92 | 8 | 7 | 0.93 | 0·34 |

CAARMS, Comprehensive Assessment of At Risk Mental State; CHR, clinical high risk; GAF, Global Assessment of Function; HC, healthy controls; NART, National Adult Reading Test; SD, standard deviation.

**Table S2. Baseline characteristics of the second sample (PROD).**

| | HC (n=12) | CHR (n=22) | HC vs CHR | | CHR-Good (n=10) | CHR-Poor (n=6) | CHR-Good vs CHR-Poor | |
|---|---|---|---|---|---|---|---|---|
| | | | T or $\chi^2$ | p | | | T or $\chi^2$ | p |
| Age in years, mean (SD) | 23·25 (4·29) | 22·55 (4·93) | 0·42 | 0·68 | 23·30 (6·26) | 23·17 (4·01) | 0·05 | 0·96 |
| Sex (male), N | 9 (75·0%) | 13 (59·1%) | 0·86 | 0·35 | 7 (70·0%) | 2 (33·3%) | 2·05 | 0·15 |
| Years of education, mean (SD) | 14·75 (1·22) | 13·14 (2·40) | 2·60 | 0·01 | 13·50 (2·37) | 12·33 (2·88) | 0·88 | 0·39 |
| Premorbid IQ estimate | 106·83 (10·13) | 106·31 (9·36) | 0·15 | 0·88 | 107·54 (10·78) | 108·90 (6·60) | -0·28 | 0·79 |
| GAF, mean (SD) | 83·17 (10·06) | 58·00 (9·85) | 7·01 | <0·001 | 60·50 (12·14) | 56·57 (7·76) | 0·69 | 0·50 |
| CAARMS Total, mean (SD) | n/a | 39·81 (12·78) | - | - | 36·30 (13·22) | 41·67 (10·93) | -0·84 | 0·42 |
| CAARMS Positive, mean (SD) | n/a | 6·68 (3·12) | - | - | 5·40 (2·84) | 8·17 (3·76) | -1·68 | 0·12 |
| CAARMS Negative, mean (SD) | n/a | 6·00 (3·15) | - | - | 5·30 (3·47) | 6·50 (3·15) | -0·69 | 0·50 |
| Antipsychotics, N | - | 1 (4·6%) | - | - | 1 (10%) | 0 | 0·64 | 0·42 |
| Antidepressants, N | - | 5 (22·7%) | - | - | 2 | 2 | 0·36 | 0·55 |
| Cannabis use, N | 8 | 15 | 0·07 | 0·79 | 6 | 3 | 0·15 | 0·70 |

CAARMS, Comprehensive Assessment of At Risk Mental State; CHR, clinical high risk; GAF, Global Assessment of Function; HC, healthy controls; NART, National Adult Reading Test; SD, standard deviation.

**Table S3. Clinic-demographic details of CHR participants with and without clinical follow-up.**

| | Sample with follow-up (n=50) | Lost to follow-up (n=17) | Sample with vs without follow-up | |
|---|---|---|---|---|
| | | | ***T* or *χ*²** | ***P*** |
| Age in years, mean (SD) | 22·97 (4·28) | 22·29 (4·14) | 0·57 | 0·57 |
| Sex (male), N | 30 (60%) | 10 (58·8%) | 0·01 | 0·93 |
| Years of education, mean (SD) | 13·82 (2·48) | 14·44 (2·26) | -0·95 | 0·35 |
| Premorbid IQ estimate | 106·91 (10·50) | 103·41 (11·33) | 1·16 | 0·25 |
| GAF, mean (SD) | 57·64 (9·67) | 60·41 (7·80) | -1·04 | 0·30 |
| CAARMS Total, mean (SD) | 41·00 (18·04) | 43·36 (19·84) | 0·43 | 0·68 |
| CAARMS Positive, mean (SD) | 8·04 (3·77) | 7·88 (2·99) | 0·16 | 0·87 |
| CAARMS Negative, mean (SD) | 5·90 (3·68) | 5·86 (3·68) | 0·04 | 0·97 |
| Antipsychotics, N | 3 (6%) | 2 (11·8%) | 0·61 | 0·44 |
| Antidepressants, N | 16 (32%) | 4 (24%) | 0·44 | 0·51 |
| Cannabis use, N | 24 (48·0%) | 10 (58·8%) | 0·60 | 0·44 |
| Global rCBF, mean (SD) | 439·54 (89·87) | 458·59 (81·94) | -0·77 | 0·44 |
| Striatal $K_i^{cer}$, mean (SD) | 0·0129 (0·001) | 0·0125 (0·001) | 0·71 | 0·20 |

Abbreviations: CAARMS, Comprehensive Assessment of At Risk Mental State; CHR, clinical high risk; GAF, Global Assessment of Function; SD, standard deviation.

**Table S4. Group differences in dopamine synthesis capacity by striatal subdivision.**

**HC (n=28) vs CHR (n=67)**

| | F | p | F* | p* |
|---|---|---|---|---|
| Associative striatum | 0·599 | 0·441 | 0·384 | 0·537 |
| Limbic striatum | 0·104 | 0·748 | 0·168 | 0·683 |
| Sensorimotor striatum | 0·602 | 0·440 | 0·376 | 0·541 |

*Adjusted for age and gender.

**CHR-Good (n=25) vs CHR-Poor (n=25)**

| | F | p | F* | p* |
|---|---|---|---|---|
| Associative striatum | 2·905 | 0·095 | 1·958 | 0·168 |
| Limbic striatum | 3·217 | 0·079 | 2·420 | 0·127 |
| Sensorimotor striatum | 0·821 | 0·372 | 0·218 | 0·643 |

*Adjusted for age and gender.

**CHR-NT (n=44) vs CHR-T (n=6)**

| | F | p | F* | p* |
|---|---|---|---|---|
| Associative striatum | 0·700 | 0·407 | 1·138 | 0·292 |
| Limbic striatum | 0·236 | 0·630 | 0·333 | 0·567 |
| Sensorimotor striatum | 0·003 | 0·960 | 0·089 | 0·767 |

*Adjusted for age and gender.

**CHR-R (n=33) vs CHR-NR (n=16)**

| | F | p | F* | p* |
|---|---|---|---|---|
| Associative striatum | 0·010 | 0·919 | 0·252 | 0·618 |
| Limbic striatum | 1·734 | 0·194 | 2·361 | 0·131 |
| Sensorimotor striatum | 0·024 | 0·878 | 0·024 | 0·878 |

*Adjusted for age and gender.

### Supplemental Figures

**Figure S1. Region-of-interest mask for the bilateral hippocampus (green) overlaid on (A) a standard brain template, and (B) a representative subject-level cerebral blood flow map in normalized space.**

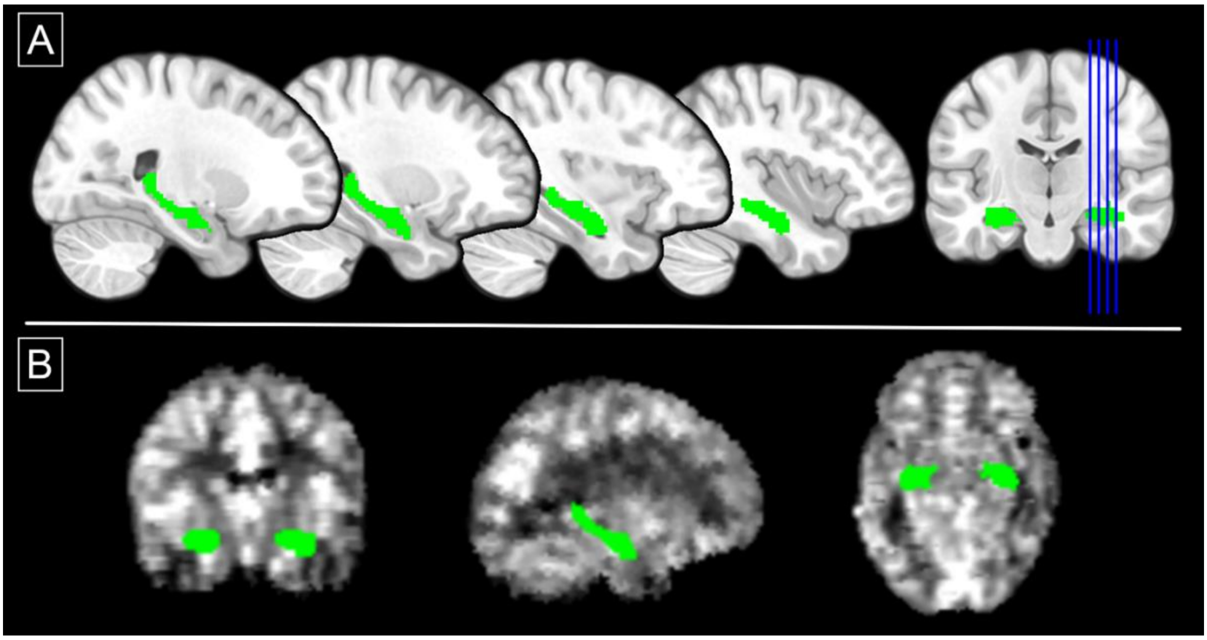

**Figure S2. Region-of-interest mask for the hippocampal subfield analysis.**

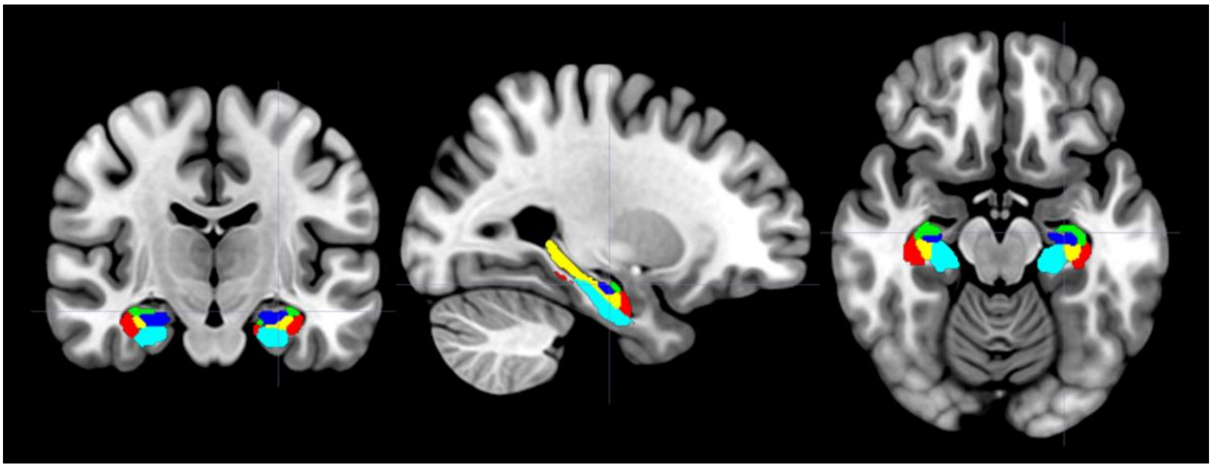

ROI masks for bilateral hippocampal subregions: dentate gyrus (yellow), subiculum (light blue), CA1 (red), CA2 (green), and CA3 (dark blue) displayed on a standard brain template.

**Figure S3. Scatterplot of follow-up GAF scores across the total CHR sample with available clinical follow-up (n=50).**

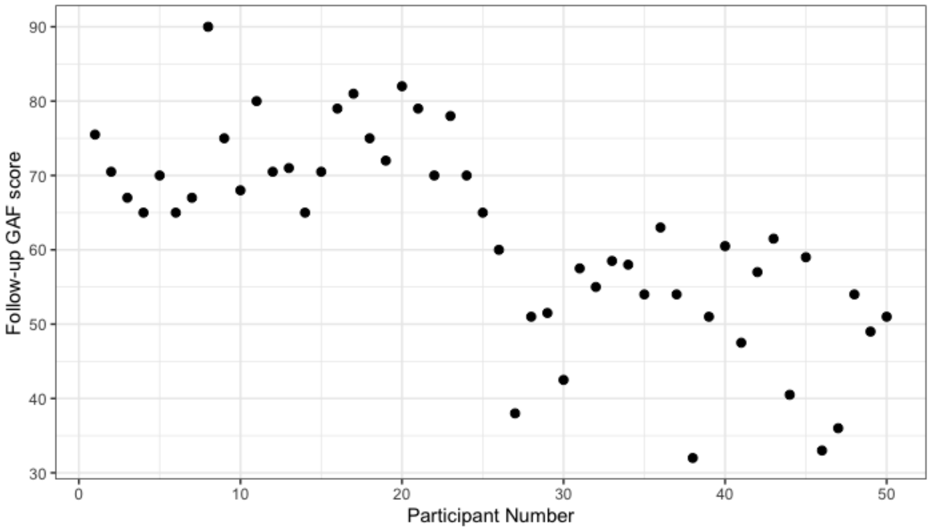

**Figure S4. Group differences in dopamine synthesis capacity (whole striatum and its subdivisions) between CHR-Poor (n=25) and CHR-Good subgroups (n=25).**

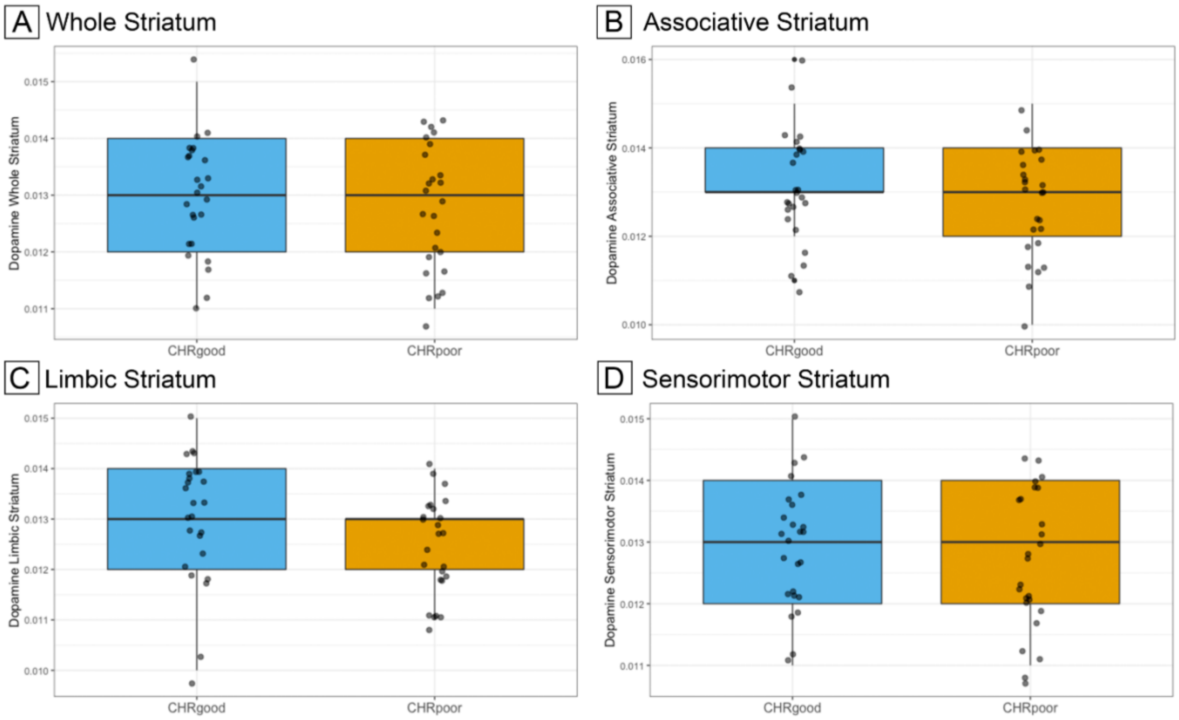

Boxplots with overlaid individual datapoints for (A) whole, (B) associative, (C) limbic, and (D) sensorimotor striatum. There were no significant differences between CHR individuals with good functional outcomes (blue) and CHR individuals with poor outcomes (orange). Striatal dopamine is quantified in $K_i^{cer}$ min$^{-1}$.

**Figure S5. Group differences in dopamine synthesis capacity (whole striatum and its subdivisions) between HC (n=28) and the total CHR sample (n=67).**

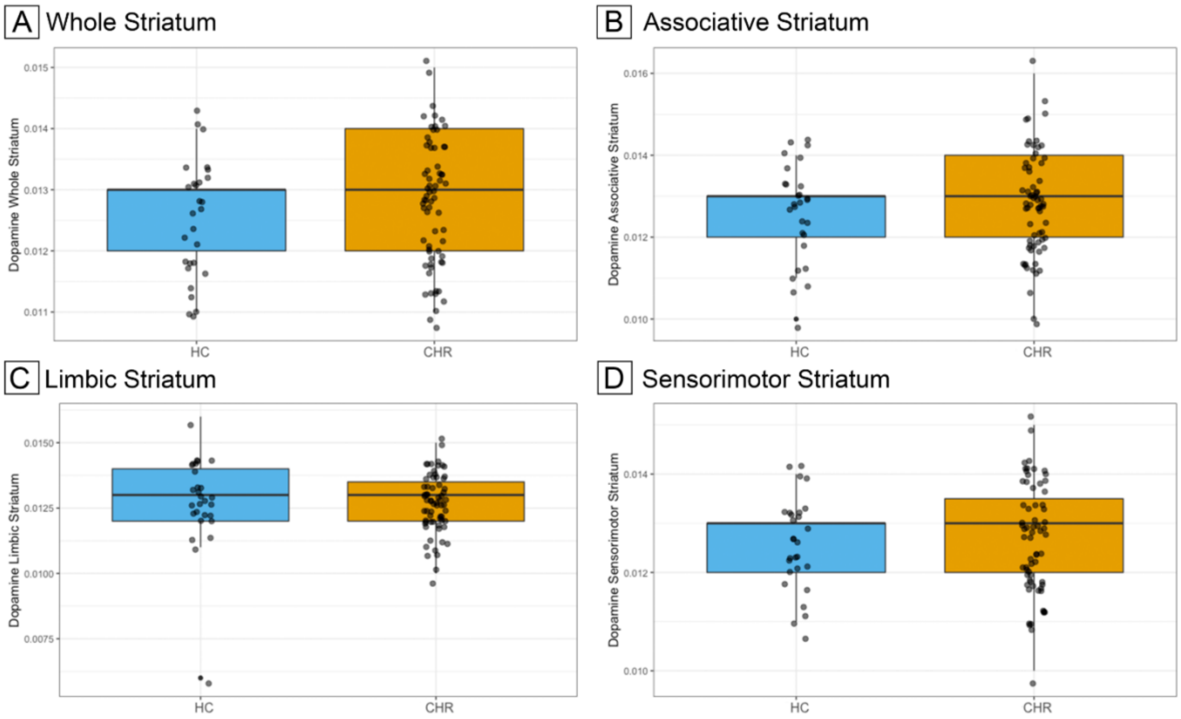

Boxplots with overlaid individual datapoints for (A) whole, (B) associative, (C) limbic, and (D) sensorimotor striatum. There were no significant differences between HC (blue) and the total CHR sample (orange). Striatal dopamine is quantified in $K_i^{cer}$ $min^{-1}$.

**Figure S6. Associations between hippocampal rCBF and dopamine synthesis capacity in striatal subdivisions in CHR individuals with poor functional outcomes compared to those with good outcomes.**

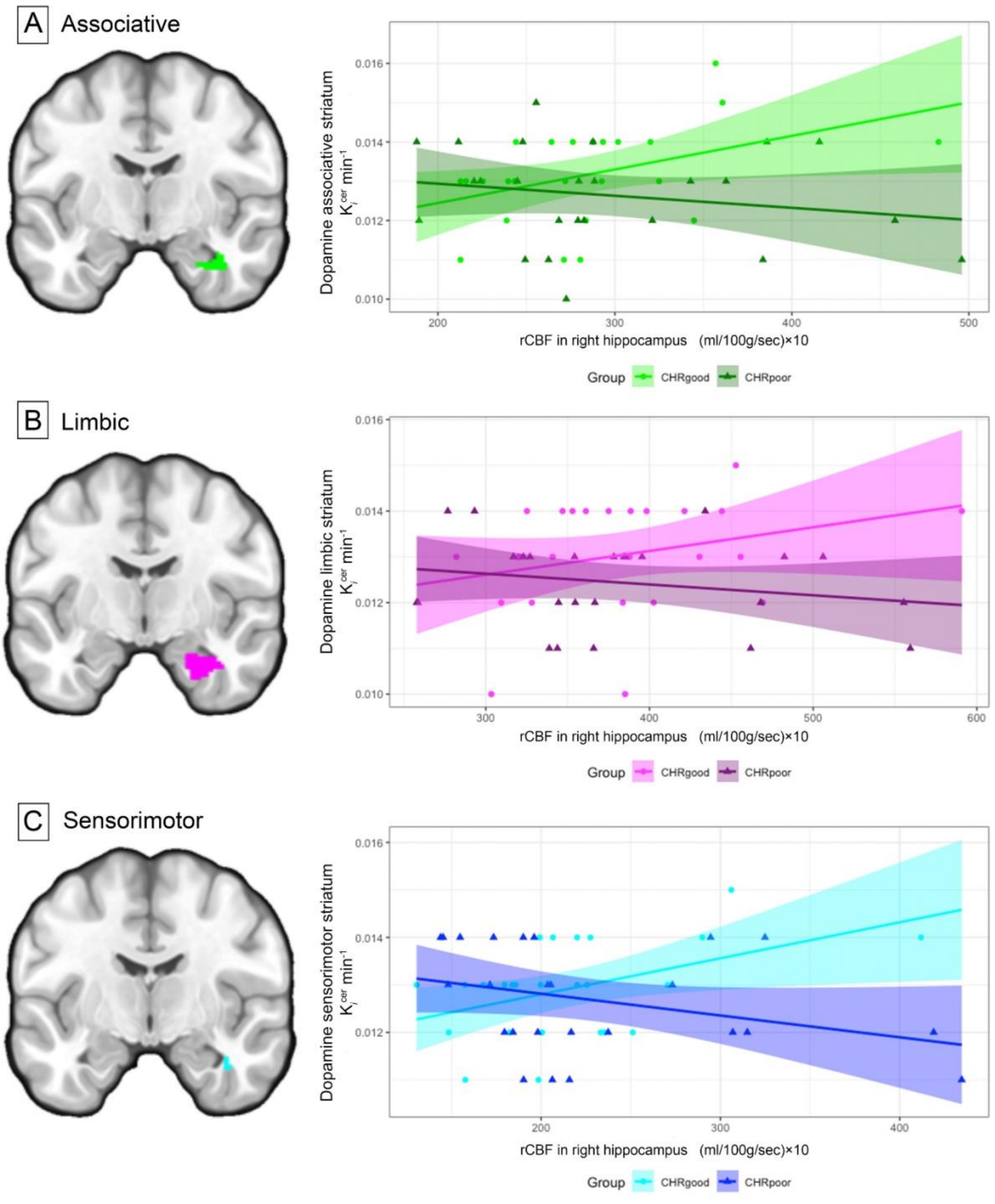

Significant hippocampal rCBF x $K_i$ interaction effects for (A) associative (pFWE=0·046), (B) sensorimotor (pFWE=0·036), and trend-level in the limbic (pFWE=0·054) striatum. Cluster-averaged effects are displayed on a standard brain template and corresponding scatterplots of the association with regression line and 95% CI.
